# Using LASSO regression to estimate the population-level impact of pneumococcal conjugate vaccines

**DOI:** 10.1101/2022.02.12.22270888

**Authors:** Anabelle Wong, Sarah C. Kramer, Marco Piccininni, Jessica L. Rohmann, Tobias Kurth, Sylvie Escolano, Ulrike Grittner, Matthieu Domenech de Cellès

## Abstract

The pneumococcal conjugate vaccines (PCVs) protect against diseases caused by *Streptococcus pneumoniae*, such as meningitis, bacteremia, and pneumonia. It is challenging to estimate their population-level impact due to the lack of a perfect control population and the subtleness of signals when the endpoint – like all-cause pneumonia – is non-specific. Here we present a new approach to estimate PCVs’ impact – using LASSO regression to predict the counterfactual outcome for vaccine impact inference. We first used a simulation study to test the performance of LASSO regression and established methods including the synthetic control (SC) approach. We found that LASSO achieved accurate and precise estimation, even in complex simulation scenarios where the association between outcome and all control variables was non-causal. We then applied LASSO to real-world data and found that it yielded estimates of vaccine impact similar to SC. The LASSO method is accurate, easily implementable, and can be applied to study the impact of PCVs and of other vaccines.

## Introduction

The bacterium *Streptococcus pneumoniae* (the pneumococcus) poses a substantial health burden globally. Although it typically colonizes the human nasopharynx asymptomatically, it can disseminate to cause a diverse array of diseases that ranges from mild (such as sinusitis and otitis media) to more severe infections (such as pneumonia) and invasive diseases (such as meningitis and septicemia)^1^. The Global Burden of Disease (GBD) study found that pneumococcal pneumonia was the most common cause of lower respiratory infection morbidity and mortality worldwide, causing 1 200 000 deaths in 2016^2^. In 2019, the GBD study also identified lower respiratory infections including pneumonia to be the leading contributor to disability-adjusted life years (DALY) among children and the elderly globally^3^.

Anti-pneumococcal vaccines were developed to combat pneumococcal infections and the most widely used ones are pneumococcal conjugate vaccines (PCVs), in which several types of the pneumococcus’ capsular polysaccharide are conjugated to carrier proteins to elicit immunity against a subset among around 100 serotypes of pneumococcus^4^. Following the widespread adoption of a 7-valent PCV (PCV7) into national childhood immunization programs, PCVs of higher valency – PCV10 and PCV13 – have been introduced^5,6^ while the third generation PCVs with even higher valency – PCV15 and PCV20 – are recently licenced^7,8^. Unlike previous anti-pneumococcal vaccines that merely reduced the risk of disease^9^, PCVs also protect against carriage of vaccine serotypes and can therefore contribute to herd immunity^10,11^.

Randomized controlled trials (RCTs) demonstrated the efficacy of PCVs against diseases (e.g. invasive diseases and, to a lesser extent, pneumonia^12^) by comparing vaccinated groups to unvaccinated groups. The efficacy measured in RCTs is different from the actual vaccine impact on a population level – that is, the reduction of disease burden in a population consisting of vaccinated and unvaccinated individuals in comparison with an otherwise similar but universally unvaccinated population^13^ – after PCV introduction. RCTs may substantially underestimate vaccine impact on the population level, because vaccinating infants with PCV protects not only the vaccinated but also unvaccinated children and adults against invasive pneumococcal diseases (IPD) and pneumonia ^14–16^.

Addressing the limitation of RCTs in vaccine impact estimation means finding a suitable unvaccinated comparison population, which is difficult, if not impossible, to identify. Hence, statistical models are routinely employed to emulate the counterfactual disease burden in a hypothetical unvaccinated version of the population^16–22^. Since statistical models rely on observational data such as the incidence rates of pneumococcal diseases before and after the introduction of PCVs, they are prone to sources of confounding (such as changes in surveillance system, reporting behavior and the demographics of the population), as acknowledged by previous studies^16,18,19,22^. Moreover, the estimation of vaccine impact on less specific endpoints, such as all-cause pneumonia, is more challenging because when multiple pathogens can contribute to the outcome, the reduction in outcome due to the change in one pathogen (the one that is targeted by the vaccine) would be much smaller^13,23,24^ and therefore can be easily missed even if there is a true effect. New methods have been developed to address these complexities; however, no consensus exists on how best to construct a counterfactual framework^21,23–26^.

In this study, we present a novel approach to construct the counterfactual for the estimation of PCVs’ impact: using Least Absolute Shrinkage and Selection Operator (LASSO) regression, which simultaneously performs variable selection and parameter estimation^27^. We first assess the performance of the LASSO regression method in a series of simulations and then apply this method to real-world hospitalization data from multiple countries. We show that LASSO regression can achieve accurate counterfactual prediction for vaccine impact inference and offer a more thorough understanding about the PCVs’ impact on pneumococcal diseases in different age-groups and in different countries.

## Methods

### Data

We used monthly hospitalization data originally published and described by Bruhn et al^23^. These data consisted of routinely collected information on reasons for hospitalization in Brazil, Chile, Ecuador, Mexico and ten states in the United States (US), provided either by the Ministry of Health or the individual countries’ health care statistics agencies. In this study, we excluded data from Brazil due to a shift in coding for the cause of hospitalization in 2008 due to a reimbursement policy change^23^. We included all available data periods (differed by country, see Table 1) for all countries except for the US – specifically, as in Bruhn et al.^23^, we excluded the period 2006 – 2010 and focused only on the early post-vaccine period. In the US dataset, counts less than 10 were masked due to privacy concerns; therefore, we imputed the masked values by randomly drawing values between 0 and 9 and further excluded the period 1994 – 1995 due to considerable masked data. The data were aggregated by the following eight age groups: 0, 1, 2 to 4, 5 to 17, 18 to 39, 40 to 64, 65 to 79, and 80+ for Ecuador, Mexico and the US. For Chile, the youngest two age groups were combined (0 to 1).

**Table 1.** The characteristics of the monthly hospitalization count datasets from Chile, Ecuador, Mexico and the US*.

| Country | Data Period | PCV Type /<br>Date of<br>Introduction<sup>57</sup> | Evaluation Period | Median monthly hospitalization<br>for all-cause pneumonia<br>from all age groups<br>(2.5<sup>th</sup>, 97.5<sup>th</sup> percentile) |
| --- | --- | --- | --- | --- |
| Chile | 2001.01 – 2012.12 | PCV10 /<br>2011.01 | 2011.07 – 2012.12 | (pre-vaccine) 5151 (2636, 12209)<br>(post-vaccine) 5036 (2745, 10092) |
| Ecuador | 2001.01 – 2012.12 | PCV10 /<br>2010.01 | 2011.01 – 2012.12 | 1958 (1085, 3230)<br>2926 (1972, 5707) |
| Mexico | 2000.01 – 2013.12 | PCV7 /<br>2006.01 | 2010.01 – 2011.12 | 2246 (1223, 4603)<br>3316 (1560, 5614) |
| US* | 1996.01 – 2005.12 | PCV7 /<br>2000.01 | 2002.01 – 2004.12 | 30089 (22045, 51035)<br>34517 (25664, 53319) |

For each country, we reported the data period included in this study, the type of PCV being introduced and the date of PCV introduction, as well as how we defined the evaluation period. We also presented the median monthly hospitalization counts with 2.5<sup>th</sup> and 97.5<sup>th</sup> percentiles for the outcome, all-cause pneumonia, from all age groups, in the pre-vaccine and post-vaccine period, to illustrate the scope of the disease burden captured in these datasets. A full description of the datasets has been published by Bruhn et al.<sup>23</sup>. (\*The US data came from 10 states: Arizona, Colorado, Iowa, Massachusetts, New Jersey, New York, Oregon, Utah, Washington, and Wisconsin.)

### Outcome variable

The primary endpoint in this study was all-cause pneumonia hospitalization, an established indicator for PCVs’ impact^16,18,19,21,23,28^. A case of all-cause pneumonia was defined as the presence of International Classification of Diseases (ICD)-10 codes J12– J18 (Chile, Ecuador and Mexico) or ICD-9 codes 480–486 (US) in the diagnostic field in the electronic hospitalization databases^23^. In the US dataset, we analyzed four additional disease endpoints: 1) IPD (pneumococcal meningitis and pneumococcal septicemia); 2) pneumococcal / lobar pneumonia; 3) all-cause pneumonia according to Griffin et al.’s definition (pneumonia listed in the first diagnostic field, or listed after a first diagnosis of sepsis, meningitis, or empyema)^29^; and 4) all-cause pneumonia with a less specific definition (pneumonia listed in any of the diagnostic fields).

### Control variables and other input variables

Following Bruhn et al.^23^, we included the time series of monthly count of control conditions (“control variables”), which may be associated with all-cause pneumonia but are not themselves affected by PCV (e.g., dermatological conditions, urinary tract infections, etc.). Counts of all control variables were log-transformed and standardized. In addition, we included 11 Fourier functions with annual periodicity to model the background seasonality of pneumonia^29,30^ over 12 months (“seasonal variables”). Finally, we included the natural logarithm of non-respiratory hospitalization as an offset to control for changes in population size or changes in the surveillance system. A complete list of all input variables can be found in Appendix 1.

### Data period

For all methods except for Interrupted Time Series (ITS), the full data period was divided into three parts:

1. the pre-vaccine period, within which the regression model was trained;
2. the implementation period, during which data were not used; and
3. the evaluation period, during which the outcome incidence rate ratio (IRR) was calculated.

The pre-vaccine period varied by country depending on the data availability and the date of vaccine introduction. The implementation period varied depending on the roll-out progress of the country. The evaluation period was at least 18 months in each country. Country-specific data periods are detailed in Table 1. The fitting period for each method is specified in Table 2.

**Table 2.** A summary of how comparator methods were implemented in this study.

| **Method** | **Variables supplied** | **Variable selection?** | **Period of fit** | **How to calculate counterfactual?** | **How to calculate IRR?** |
| --- | --- | --- | --- | --- | --- |
| **ITS** | 11 seasonal variables, intervention indicator variable | No | Whole period | By predicting post-vaccine period outcome after changing indicator variable from “1” to “0” | Fitted outcome / Counterfactual outcome |
| **LASSO** | 11 seasonal variables, control variables | Yes | Pre-vaccine period | By predicting post-vaccine period outcome based on pre-vaccine fit | Observed outcome / Counterfactual outcome |
| **SC** | 11 seasonal variables, control variables | Yes | Pre-vaccine period | By predicting post-vaccine period outcome based on pre-vaccine fit | Observed outcome / Counterfactual outcome |
| **STL+ PCA** | 11 seasonal variables, control variables | No* | Pre-vaccine period | By predicting post-vaccine period outcome based on pre-vaccine fit | Observed outcome / Counterfactual outcome |

\*STL+PCA does not select variables but selects a principal component that summarizes the extracted smoothed trends of variables supplied.

### Statistical model – LASSO regression

LASSO is an extension of linear regression that decreases the variance of regression coefficients and the prediction error by adding a term to the log-likelihood to penalize the complexity of the model^27^. This leads to a parsimonious model with a subset of control variables that best predicts the outcome. To estimate the penalty parameter, we first generated a grid of 100 values for the penalty parameter and fitted LASSO regression to the pre-vaccine period data for each value in the grid. Next, we selected the best value for the penalty using either 10-fold cross validation (CV) or Akaike Information Criterion (AIC)^27^. In a 10-fold CV, the pre-vaccine data period was randomly divided into 10 groups of equal size, with 9 groups forming the training set and 1 group forming the test set. A model was fit on the training set and the minimized mean squared error (MSE) was obtained when tested on the test set. This was repeated 10 times to yield an average MSE. This was repeated 100 times on each value in the grid of penalty parameter and the penalty with the lowest MSE was selected. Using the AIC for the penalty selection, we fitted LASSO regression to the pre-vaccine data period and the penalty with the lowest AIC was selected.

We tested two variants of the LASSO regression model: the first one included all seasonal variables by default (season-forced, SF); the second one treated seasonal variables as control variables and allowed LASSO regression to select from them (season-unforced, SU). The selected model was re-fitted onto the entire pre-vaccine period to predict the counterfactual outcome 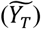 during the evaluation period – that is, the hospitalization counts that would have occurred in the population if PCV had not been introduced, assuming the distribution and associations of the population features captured in the pre-vaccine period data remained unchanged. With the LASSO-predicted counterfactual under the no-vaccine scenario and the observed outcome (*Y*_*T*_), we calculated the vaccine impact using Equation 1. An IRR less than 1 indicates a reduction in all-cause pneumonia hospitalization due to the vaccination program.

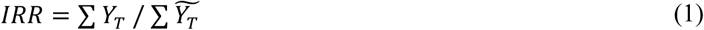

where *T* is the set of time points during the evaluation period.

### Statistical model – other methods

We compared LASSO regression to three established methods in the field of vaccine impact estimation. The key features of the implementation of LASSO regression and all comparator methods are summarized in Table 2. The three comparator methods are described below:

1. Interrupted Time Series (ITS) is a method that includes an indicator variable for vaccination, secular trends before and after PCV introduction, and background seasonality^16,32–34^. Following the procedures described in Bernal et al.^33^, we applied a standard ITS with a Poisson model that contained the date, PCV’s availability coded as an indicator variable (0 = no PCV, 1 = PCV in place), PCV’s continuous effect coded as the time elapsed from PCV introduction, and all the seasonal variables as covariates, with the logarithm of non-respiratory hospitalization as the offset. The model was fitted to the whole period of data and the vaccine impact was calculated using Equation 2:

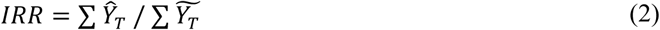

where *ŷ*_*T*_ is the fitted outcome and 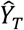 is the counterfactual outcome during the evaluation period. We used the fitted model to predict the counterfactual outcome 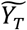 during the evaluation period in the absence of vaccination (i.e., indicator variable set to “0” for all time points).
2. In accordance with the synthetic control (SC) method^23,25^, time series of different control variables were weighted according to their fit to the outcome time series in the pre-vaccine period using Bayesian variable selection. The weighted time series were jointly used to predict the counterfactual outcome 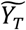. The model was adjusted for background seasonality using 11 monthly indicator variables and the logarithm of non-respiratory hospitalization was included as a covariate. The vaccine impact was calculated using Equation 1.
3. For the Seasonal-Trend decomposition using LOESS plus Principal Components Analysis (STL+PCA) method, a smoothed trend for each of the control variable’s time-series was extracted with seasonal-trend decomposition using locally-estimated scatterplot smoothing (LOESS)^24^. A PCA was performed on the extracted smoothed trends, and the first principal component was selected as the composite trend, which was used as a covariate in a regression model to predict the counterfactual outcome 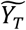. The vaccine impact was calculated using Equation 1.

### Performance assessment with simulated data

To assess the performance of all methods to estimate vaccine impact, we designed a simulation study. We generated the outcome, monthly pneumonia hospitalization (*Y*_*t*_), based on a combination of *n* (*n = {5, 10}*) control variables (*X*_1_, *X*_2_, …, *X*_*n*_) randomly selected from the list of control variables available in the Mexico dataset^23^. We then incorporated an intercept (*α*), the logarithm of non-respiratory hospitalization as the offset (ln(*NRH*_*t*_)), background seasonality (*S*_*t*_), and a vaccine impact component (*γ*) into the equation to generate the logarithm of the expected number of monthly pneumonia hospitalization. A pre-determined value was assigned to *γ* starting from the time point of PCV introduction (*t*_*vac*_). Assuming a Poisson distribution for the outcome, we simulated 100 time series; thus, the variability of the simulated time series originated from the Poisson variation. The model is represented by Equation 3:

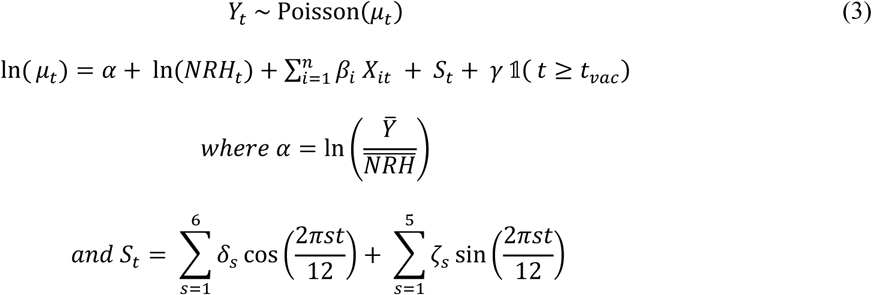

The intercept, α, is calculated as the logarithm of the mean ratio of pneumonia hospitalization to all non-respiratory hospitalization 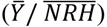. For the association of any control variable and the outcome not to be unrealistically strong, the values assigned to the *β* of the included control variables were randomly sampled from a uniform distribution with range –0.3 to 0.3, such that a change of one standard-deviation in the control variable, holding the other variables constant, would result in 0.74 – 1.35-fold change in the outcome. *S*_*t*_ was modelled as a Fourier series of 11 terms that consisted of 6 cosine and 5 sine functions. We assigned a value of 0.5 to the *δ* of the first cosine function such that the outcome peaked in January and oscillated approximately 50% above and below the annual mean to mimic pneumonia seasonality in the real world. The impact of vaccination was modeled by the parameter *γ*. In all simulations, we assumed a vaccine with null impact (*γ* = 0), IRR=1. The simulated data were screened to be realistic, such that the maximum ratio of annual maximum-to-minimum for the expected count of the outcome in any simulation set would not exceed 10.

We tested the performance of all methods in four different scenario types. First, we used five randomly selected control variables and a seasonal variable to simulate the outcome. This analysis was performed five times (simulation sets A to E) with re-sampling of the 5 control variables. To illustrate this process using an example, we plotted the first set (set A) of five control variables and their respective assigned *β* values used to generate the outcome time-series in Figure 1A and the generated outcome time series in Figure 1B. Second, we repeated this analysis with 10 control variables (simulation sets F to J). Third, we tested the performance of all methods on sparse data, which may affect the performance of these methods, as has been previously suggested^24^. We generated the sparse data (simulation set K) by taking a 10% binomial sub-sample of the first set (set A) of simulated data. A flow-chart for the outcome simulation procedures can be found in Appendix 2 Figure S1; plots similar to Figures 1A and 1B for simulation sets A to E can be found in Appendix 2 Figure S2. The complete list of control variables used to simulate the outcome can be found in Appendix 1.

**Figure 1.**
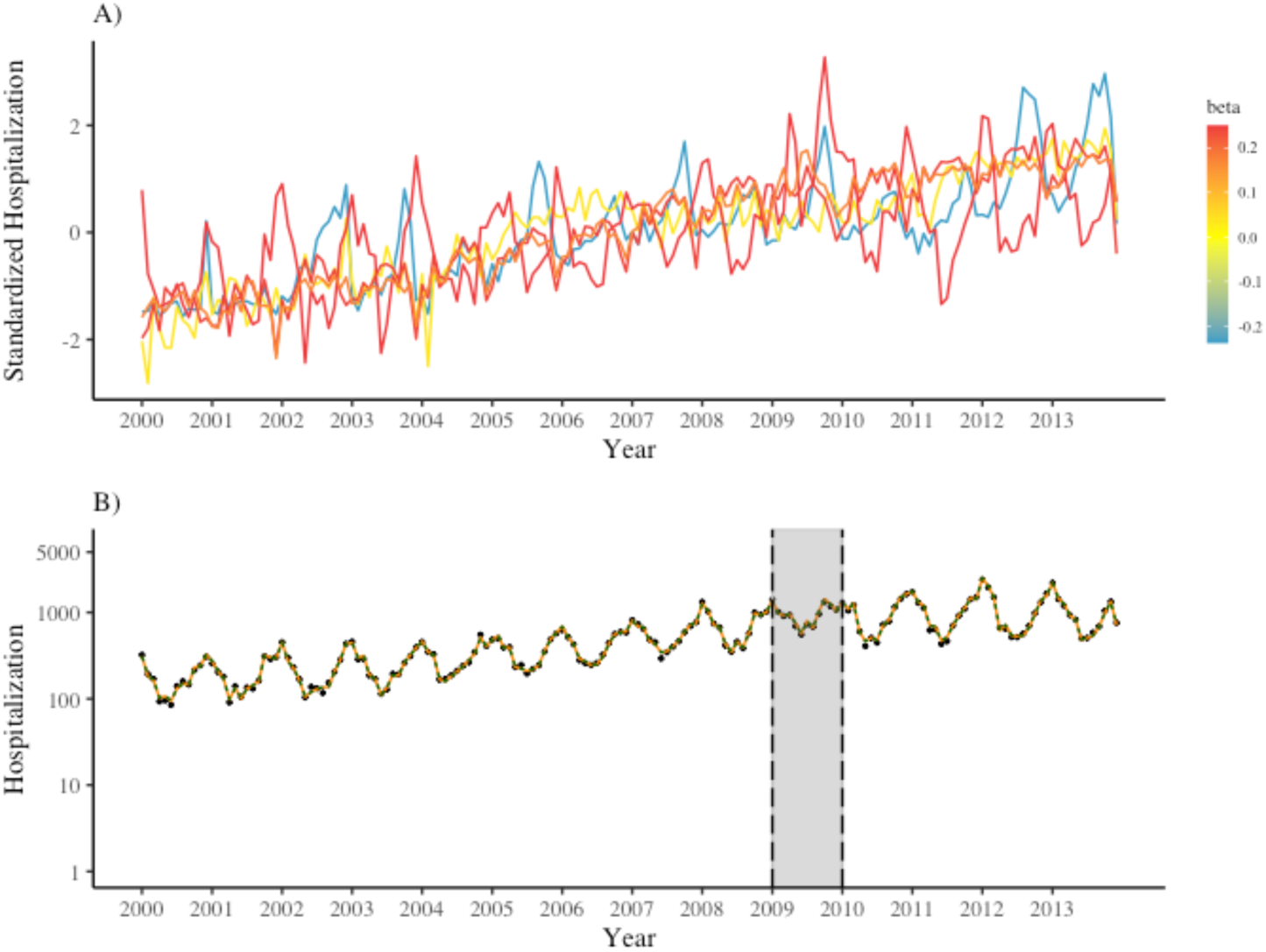
Simulation of outcome time series based on five randomly selected control conditions. A) Time series of the first set of five control variables selected randomly for the simulation of the outcome data, colored by the association size, *β*, assigned to each of them (see Supplementary Material 2 for other scenarios that used different sets of five control variables to generate the outcome). The five control variables shown here are admission due to health exams (orange, *β* = 0.17), non-pneumonia infection (blue, *β* = -0.23), dermatological condition (red, *β* = 0.24), non-pneumococcal septicemia (yellow, *β* = 0.04), and e) bronchitis and bronchiolitis (red, *β* = 0.25). B) Time series of the simulated outcome, all-cause pneumonia (black point) and the model-prediction from LASSO-SF (orange solid line) and LASSO-SU (green dotted line). The implementation period is marked by the grey area.

Finally, we tested all methods in a fourth scenario type (simulation sets L to P), in which the variables causing the outcome were not available. We used three control variables (C1, C2 and C3) to generate four conditions (Z1 to Z4) and the outcome, such that the conditions Z1 to Z4 and the outcome were not causally related but non-causally associated via common causes. We then removed C1, C2 and C3 together with their associated control variables (that is, diagnosis in the same ICD-10 chapter) from the list of control variables. Such a framework is more realistic because observed associations between different causes of hospitalization is likely due to common causes rather than direct causal influence. The Directed Acyclic Graph (DAG) for the underlying data generation process under this framework can be found in Appendix 2 Figure S3 and the control variables used for outcome simulation can be found in Appendix 1.

We evaluated each method’s performance by comparing each method’s estimates to the true IRR, which was 1. IRR estimates less than 1 indicated an overestimation of vaccine impact; IRR estimates greater than 1 indicated an underestimation of vaccine impact. In the LASSO regression and ITS, we report uncertainty of estimation as the 95% prediction intervals (95% PI) of the IRR obtained from each simulation, extracted from the 2.5^th^ and 97.5^th^ percentiles of the Poisson distribution of the predicted value. In SC and STL+PCA, we report the 95% credible intervals (95% CI) of the IRR from each simulation, extracted from the 2.5^th^ and 97.5^th^ percentiles of the Bayesian posterior distributions. Currently, the calculation of standard error remains an unresolved challenge in frequentist LASSO methods and the usual statistical constructs such as confidence intervals and p-values do not exist in its implementation^35,36^. Therefore, we reported different uncertainty measures for different methods, and they are not directly comparable. For each set of 100 simulations, we measured the accuracy of each method by calculating the mean IRR with standard deviation (SD). We measured the precision as the width of the 95% uncertainty intervals (95% UI). For each set of 100 simulations, we assessed performance stability by coverage (proportion of the time that the uncertainty intervals contain the true value). For each scenario type, we report these performance indicators as a range.

### Application to real-world data

In the analysis on the primary endpoint (all-cause pneumonia), the two variants of LASSO regression, LASSO-SF and LASSO-SU, as well as SC, were applied to each age group in each country. Similar to the procedure used on the simulated data, we fitted models to the pre-vaccine period and used these fitted models to predict the counterfactual outcomes. We performed model selection, the prediction of the counterfactuals, and the calculation of vaccine impact using the same procedures in the performance assessment using the simulated data; we then compared the results from the three methods.

We then applied LASSO-SF, LASSO-SU and SC on the US data using different endpoints (IPD, pneumococcal pneumonia, and two definitions of all-cause pneumonia) and compared the results. We performed a sensitivity analysis by removing “bronchitis and bronchiolitis” from the list of possible control variables for LASSO selection, because this control variable could be affected by PCV and violate the assumption that all control variables are not affected by the health intervention, by definition^23,37^.

### Numerical implementation

All analyses were conducted in RStudio with R version 4.1.0 (R)^38^. LASSO regression was implemented using the package “glmnet” version 4.1-2^39^. SC and STL+PCA were implemented using the package “InterventionEvaluatR” version 0.1^40^. The project’s R dependencies were recorded by the package “renv” version 0.14.0^41^ for reproducibility.

## Results

### Performance assessment with 5 causal control variables

We present the vaccine impact estimated by each method for each simulation in five scenarios (sets A to E) in Figure 2. In each scenario, a different set of five randomly selected control variables was used to generate the outcome. LASSO-SF and LASSO-SU performed well in all five scenarios; both achieved high coverage (SF: 97–100%; SU: 96–100%) and accurate mean IRR (SF: 1.00–1.02; SU: 1.00–1.03) with good precision (SF; SU: 0.12–0.13), as shown in Figure 2 and Table 2. The estimates obtained by LASSO-SF and LASSO-SU were similar. In general, LASSO regression tended to select the causal variables (Appendix 3 Figure S4). When comparing CV selection and AIC selection, we did not observe different performance in terms of accuracy and precision, but we noticed CV selection resulted in models with more variables while AIC selection led to more parsimonious models (Appendix 3 Figure S5).

**Figure 2.**
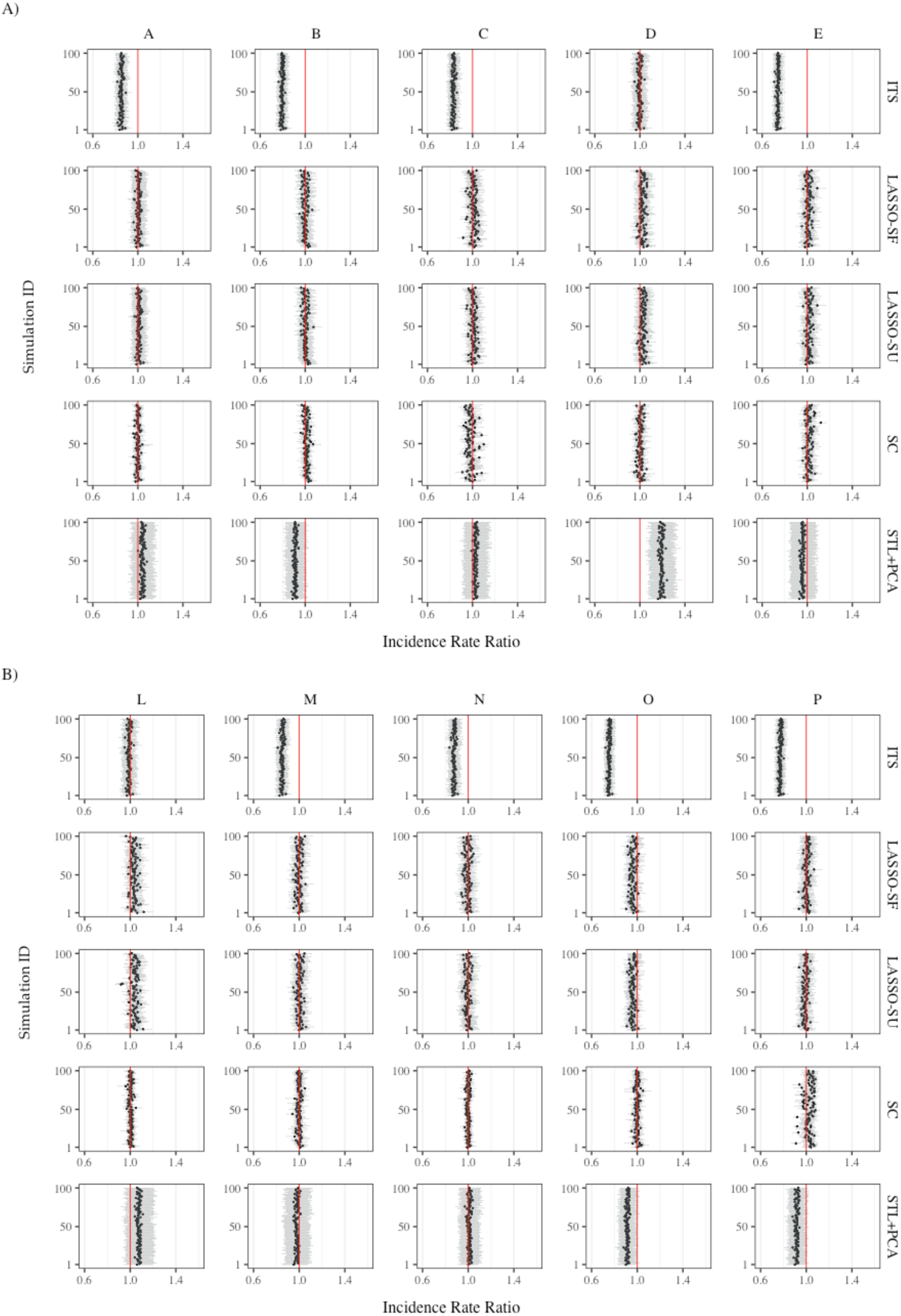
Incidence rate ratios estimated by various methods for simulation 1 to 100 in different simulation scenarios. A) Each row shows the estimates using a different method, from top to bottom: ITS, LASSO-SF, LASSO-SU, SC, and STL+PCA. Each column represents a scenario with outcome simulated with a different set of five causal control variables and one seasonal variable, from left to right: set A – health exams, bronchitis and bronchiolitis, dermatological condition, non-pneumonia infection, and non-pneumococcal septicemia; set B – non-pneumococcal septicemia, urinary tract infection (UTI), diabetes, stroke, and injury; set C – human immunodeficiency virus (HIV) infection, cholelithiasis, dermatological condition, health exams, diabetes, and bronchitis and bronchiolitis; set D – UTI, dermatological conditions, health exams, diabetes, and bronchitis and bronchiolitis; and set E – HIV infection, gynecological conditions, gastrointestinal conditions, non-pneumococcal septicemia, and appendicitis. Each panel shows the result in 100 simulations, the points represent the estimated Incidence Rate Ratio (IRR) and the error bars represent the 95% uncertainty intervals. The red vertical line indicates the true vaccine impact in the simulation, which is 1 in all of our simulation scenarios; here, an IRR larger than 1 means underestimation of vaccine impact and an IRR lower than 1 means overestimation of vaccine impact. B) Similar to A), Each row shows the estimates using a different method. Each column represents a scenario with outcome simulated from a different set of three causal control variables which were then removed alongside other control variables under the same ICD chapter, leaving behind only non-causal control variables. From left to right: set L – neoplasm, UTI, and bronchitis and bronchiolitis; set M – neoplasm, UTI, and diabetes; set N – health exams, UTI, and bronchitis and bronchiolitis; set O – health exams, UTI, and diabetes; and set P – gastrointestinal conditions, UTI, and diabetes.

Other methods showed variable performance. ITS yielded accurate and precise estimates with high coverage in one scenario (set D) but the estimates were biased, although precise in the other scenarios (sets A, B, C & E), resulting in variable coverage (0–100%). Similarly, the mean IRR estimated by STL+PCA was biased in some scenarios (sets B & D), causing the coverage to be variable (0–100%). SC showed relatively high coverage (78– 94%) and accurate mean IRR (0.99–1.02) with good precision (0.07–0.11). The performance indicators of all methods are summarized in Table 3.

**Table 3.** Performance of the five methods in estimating vaccine impact under different simulation scenarios.

| Method | Range of Mean IRR*<br>(Range of SD*) | Range of Mean Width of<br>95% Uncertainty Intervals* | Range of<br>Coverage* |
| --- | --- | --- | --- |
| Simulation sets A-E (outcome simulated using 5 causal control variables) |  |  |  |
| ITS | 0.74 – 1.00 (0.010 – 0.015) | 0.08 – 0.12 | 0 – 100% |
| LASSO-SF | 1.00 – 1.02 (0.015 – 0.026) | 0.12 – 0.13 | 97 – 100% |
| LASSO-SU | 1.00 – 1.03 (0.014 – 0.023) | 0.12 – 0.13 | 96 – 100% |
| SC | 0.99 – 1.02 (0.014 – 0.035) | 0.07 – 0.11 | 78 – 94% |
| STL+PCA | 0.91 – 1.19 (0.012 – 0.015) | 0.17 – 0.23 | 0 – 100% |
| Simulation sets F-J (outcome simulated using 10 causal control variables) |  |  |  |
| ITS | 0.75 – 1.15 (0.009 – 0.017) | 0.09 – 0.16 | 0 – 0% |
| LASSO-SF | 0.99 – 1.03 (0.020 – 0.028) | 0.14 – 0.14 | 96 – 100% |
| LASSO-SU | 0.99 – 1.03 (0.020 – 0.027) | 0.14 – 0.14 | 91 – 100% |
| SC | 0.99 – 1.01 (0.023 – 0.040) | 0.08 – 0.13 | 87 – 95% |
| STL+PCA | 0.87 – 1.23 (0.010 – 0.017) | 0.19 – 0.23 | 0 – 100% |
| Simulation set K (10% as sparse as set A) |  |  |  |
| ITS | 0.86 (0.041) | 0.31 | 74% |
| LASSO-SF | 1.02 (0.044) | 0.40 | 100% |
| LASSO-SU | 1.03 (0.042) | 0.41 | 100% |
| SC | 1.02 (0.046) | 0.23 | 98% |
| STL+PCA | 1.03 (0.045) | 0.25 | 97% |
| Simulation sets L-P (all causal control variables removed) |  |  |  |
| ITS | 0.75 – 0.99 (0.011 – 0.014) | 0.08 – 0.12 | 0 – 100% |
| LASSO-SF | 0.96 – 1.03 (0.021 – 0.027) | 0.11 – 0.13 | 79 – 100% |
| LASSO-SU | 0.96 – 1.04 (0.019 – 0.030) | 0.11 – 0.13 | 81 – 100% |
| SC | 1.00 – 1.02 (0.012 – 0.036) | 0.06 – 0.11 | 78 – 97% |
| STL+PCA | 0.91 – 1.08 (0.011 – 0.014) | 0.15 – 0.24 | 16 – 100% |

We applied five methods (ITS, LASSO-SF, LASSO-SU, SC, and STL+PCA) to sixteen sets of 100 simulations. The sixteen sets can be grouped into four types of simulation scenarios: sets A to E – outcome simulated using 5 causal control variables; sets F to J – outcome simulated using 10 causal control variables; set K – same as set A but outcome count reduced to 10% as sparse; and sets L to P – outcome simulated using 3 causal control variables, which were then removed. We summarized the Incidence Rate Ratio (IRR) estimated for each of the 100 simulations using the mean IRR with standard deviation (SD). We summarized the precision of the IRR estimated for each of the 100 simulations using the mean width of 95% uncertainty intervals. We report the range of mean IRR and the range of SD, as well as the range of mean width mean width of 95% uncertainty intervals, for each type of simulation scenarios, except for set K, there was only one set of simulation and therefore we report the mean IRR with SD, mean width or 95% uncertainty intervals, and coverage, instead of a range\*.

### Performance assessment with 10 causal control variables

The performance of LASSO-SF and LASSO-SU remained robust in another five scenarios (sets F to J), in which different sets of ten randomly selected control variables were used to generate the outcome. As the causal control variables increased from 5 to 10, the number of control variables that were consistently selected by LASSO-SF and LASSO-SU also increased (Appendix 4 Figure S7). Again, the performance of SC was satisfactory and consistent, while that of other methods appeared to be variable (Appendix 4 Figure S6). The performance is summarized in Table 3.

### Performance assessment with sparse data

When the monthly hospitalization counts became as sparse as 10% of the first simulation scenario (set K), the performance of all methods remained consistent in terms of accuracy, but the precision notably decreased as the 95% UIs of the IRR estimated by all methods widened considerably, which in turn increased coverage.

### Performance assessment with non-causal control variables

When tested on the data generated from three causal control variables (C1, C2 and C3) that were subsequently removed (sets L to P), the estimation by LASSO-SF and LASSO-SU remained accurate and precise (Table 3, Figure 2). In absence of C1, C2 and C3, LASSO-SF and LASSO-SU selected the control variables that were associated with the outcome via the common parent, such as Z2, Z3 or Z4. We also observed that LASSO-SF and LASSO-SU preferentially selected the control variable more strongly associated with the common parent, Z2; whereas Z1, the control variable with a weaker association with the common parent, was almost never selected in all five scenarios (Appendix 4 Figure S8).

### Application to real-world data

**T**he characteristics of the four country’s datasets used in this analysis are summarized in Table 1. The IRR estimated by LASSO-SF and LASSO-SU were comparable to those by SC for Chile, Ecuador, Mexico and the US. The three methods generally arrived at the same conclusion as to whether there was a significant impact of PCV, except for two instances: (1) the two LASSO methods found a significant impact of PCV in the age group 40 to 64 years in Chile, in age groups 18 to 64 years in Ecuador, and in the age group 40 to 64 years in the US while SC did not, and (2) the SC method detected a significant impact of PCV in the age groups 0 to 1 year in Mexico, which was not detected by the two LASSO methods. Complete results are shown in Figure 3. In general, we found that LASSO delivered comparable estimates to SC, which has been shown to be a reliable method in vaccine impact estimation^23,25^.

**Figure 3.**
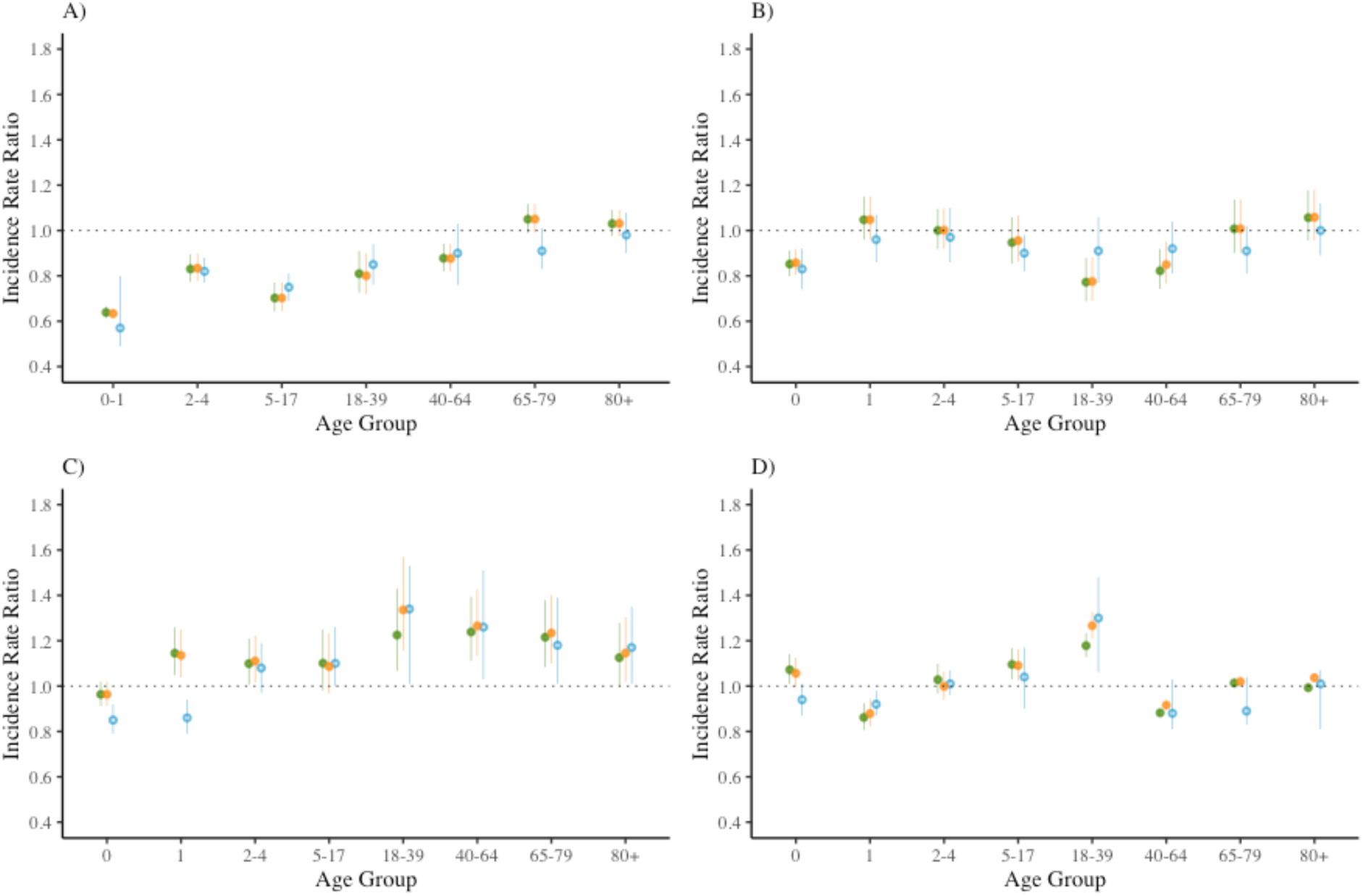
Age-group-specific incidence rate ratios (IRR) in four countries, estimated by two LASSO methods and SC. A – D) Each panel shows the age-group-specific incidence rate ratios (IRR) for all-cause pneumonia in a population whose infants were vaccinated with pneumococcal conjugate vaccines (PCV) compared to a counterfactual population in which PCV was never introduced, estimated by LASSO-SF (green), LASSO-SU (orange), and SC (blue). The four countries are A) Chile; B) Ecuador; C) Mexico; and D) the US. The 95% prediction intervals (PI) of estimates by LASSO-SF and LASSO-SU are shown by the error bars joint at a filled circle; the 95% credible intervals (CI) of estimates by SC are shown by the error bars joint at an open circle. 95% PI and 95%CI are different uncertainty measures and they are not directly comparable.

The results using Ecuador, Mexico, and US data were sensitive to removing “bronchitis and bronchiolitis” from the list of control variables that LASSO regression and SC could choose from. In Ecuador, the reduction in all-cause pneumonia hospitalization attenuated in the youngest age group, but remained statistically significant in age groups 18 to 64 years. In Mexico, the reduction in the youngest two age groups detected by SC was also attenuated and was no longer statistically significant. In the US, only a marginal reduction was detected by LASSO-SF and LASSO-SU in the age group 40 to 64 years and not in older adults before removing “bronchitis and bronchiolitis”, but after doing so, a more pronounced reduction was observed in all the age groups from 18 to 79 years. Full results are shown in Appendix 5 Figure S9.

Finally, we applied LASSO-SF, LASSO-SU and SC on IPD hospitalization data in the US and compared the results using four different endpoints: IPD, pneumococcal pneumonia, all-cause pneumonia (more specific definition), and all-cause pneumonia (more inclusive definition). LASSO-SF found a statistically significant reduction in IPD hospitalization across all age groups, except for the age group 5 to 17 years. LASSO-SU and SC found a significant reduction in IPD hospitalization in age groups younger than 5 years and older than 64 years. As the endpoint definition became less specific, the reduction point estimate became smaller in size and the 95% UI more often included 1. The results are shown in Figure 4.

**Figure 4.**
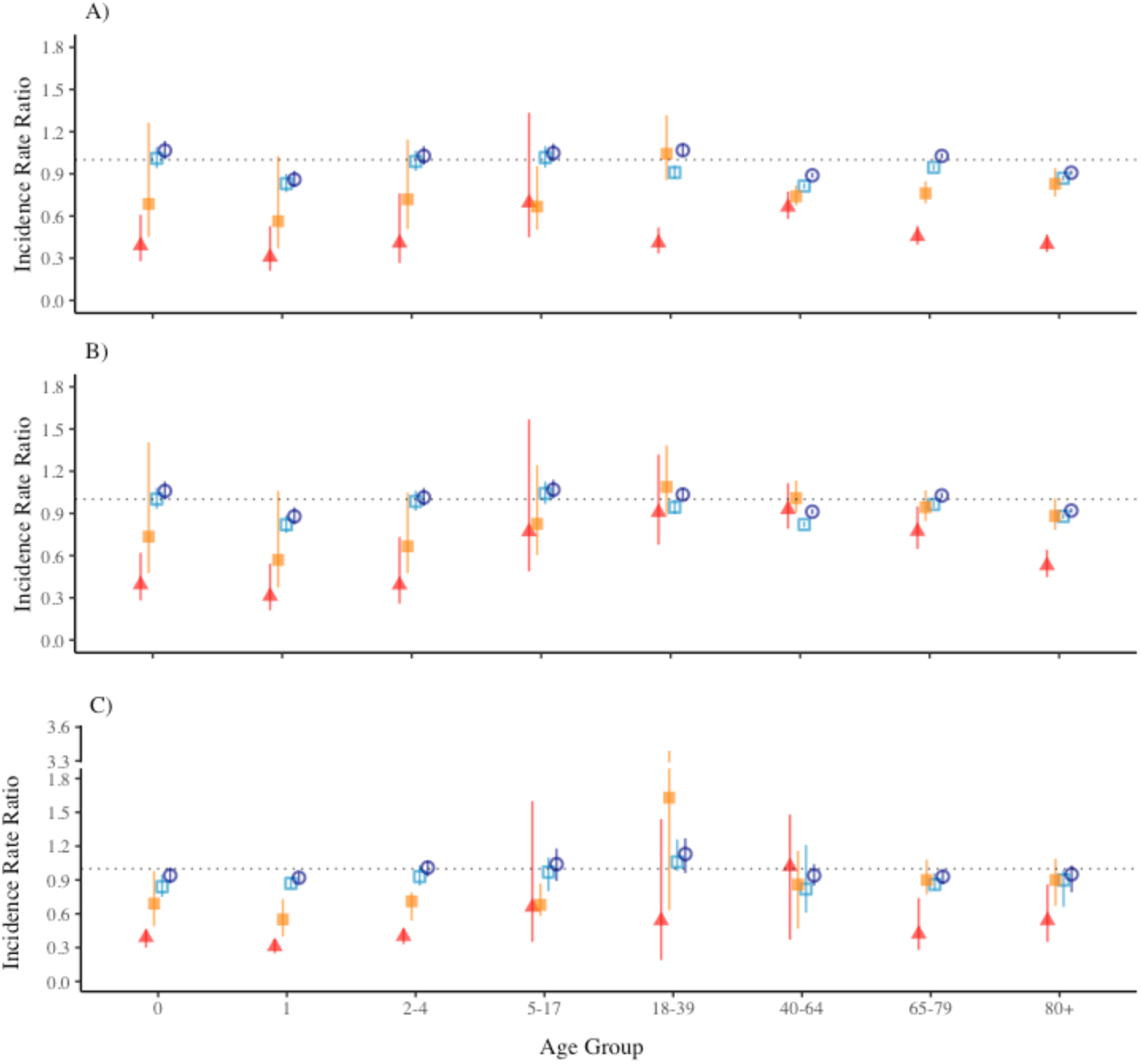
Age-group-specific incidence rate ratios (IRR) regarding invasive pneumococcal diseases (IPD) and other disease endpoints in the US, estimated by two LASSO methods and SC. A – C) Each panel shows the age-group-specific incidence rate ratios (IRR) for four diseases endpoints in a population whose infants were vaccinated with pneumococcal conjugate vaccines (PCV) compared to a counterfactual population in which PCV was never introduced, estimated by A) LASSO-SF, B) LASSO-SU, and C) SC. The four endpoints used were invasive pneumococcal diseases (IPD) (red filled triangle), pneumococcal pneumonia (orange filled square), all-cause pneumonia as primary diagnosis or as first diagnosis after sepsis, meningitis, or empyema (light blue open square), and all-cause pneumonia as primary or non-primary diagnosis (dark blue open circle). The error bars represent the A & B) 95% prediction intervals (PI) and C) 95% credible intervals (CI) of the IRR estimates. 95% PI and 95%CI are different uncertainty measures and they are not directly comparable.

## Discussion

In this study, we aimed to assess whether LASSO regression models can accurately estimate vaccine impact. Using a simulation study, we first assessed the performance of LASSO regression compared with other commonly-implemented methods, such as ITS, SC^23,25^ and STL+PCA^24^. We then applied LASSO regression and SC^23,25^ to real-world data and compared their results. Overall, we found that LASSO regression allowed for accurate and precise estimation of vaccine impact and performs comparably to established methods, such as SC^23,25^.

The results from the simulation study showed that LASSO regression was able to estimate the pre-determined vaccine impact accurately and precisely, and its performance remained stable even under more complex data simulation procedures, such as the one without any causal variables. While ITS was able to estimate the pre-determined vaccine impact accurately in some simulation scenarios, its performance was not robust across scenarios. As ITS did not include any control variables but only the offset and seasonal terms, its assumption that the characteristics in the population remained unchanged throughout the study period limited its performance^42^. In practice, control variables can be included in more advanced ITS models to improve performance^16,42^; however, the process of hand-picking control variables is subjective and can introduce biases into the analysis^43^. By design, ITS assumes a linear (or exponential, when a log-link function is used) trend for the continuous effect of an intervention^23,42,44^. This assumption can limit the method’s validity because the nature of continuous effect of an intervention is often unknown or difficult to ascertain.

We compared LASSO regression with two other methods – the SC approach^23,25^, and STL+PCA^24^. In contrast to *a priori* selection of control conditions^21^, these methods use data-driven approaches to select various control conditions to generate the counterfactual comparator. The advantage of relying on the observed pattern of associations of several conditions is to reduce confounding of the causal relationship of interest even when confounders are unknown or when the network of causal pathways are complicated^21,23,24^.

The results from the simulation study showed stable performance of LASSO methods and SC, while the performance of STL+PCA was not robust. One possible explanation for the biased estimation by STL+PCA in some of the simulation scenarios is that only one principal component was used for the counterfactual prediction, which may not be sufficient in some of the simulation scenarios. As shown in the stimulation study, LASSO regression had the tendency to select the causal variables, or the associated variables when causal variables were not available, which is consistent with its feature in identifying few predictors with strong association^27,45^.

When we applied LASSO regression to the real-world data, we found that PCV statistically significantly reduced all-cause pneumonia hospitalization in the youngest children age groups in Chile and Ecuador, which is consistent with existing literature on PCV impact in Chile^46,47^ and Ecuador^37,48^, but we did not observe similar results in the youngest age groups in Mexico and in the US. This is in contrast to the established PCV impact estimated among children in Mexico^48^ and the US^29^. Although we found a significant reduction in pneumonia hospitalization in adult age groups (18 to 64 years) in Chile and Ecuador, we did not observe similar results in Mexico and the US. This is in contrast to the indirect impact in older age groups in different countries summarized by Tsaban and Ben-Shimol in a systematic review^49^. Part of these discrepancies may be explained by the pragmatic aspects of vaccine roll-out and by the choice of evaluation period. In Mexico, PCV7 was first given to children in the resource-poor regions in 2006 before its official introduction in 2008^50^; therefore, the pneumonia hospitalization from 2006 to 2008 might have experienced a reduction already due to partial PCV7 roll-out. The initial universal vaccination in Mexico only covered 2 doses instead of 3 doses in 2008 and 2009 due to financial constraints^50^, which could affect PCV’s indirect impact in older age groups. In the US, initial low PCV 3-dose coverage during 2002 – 2004 may have been insufficient to reach herd immunity in the older age groups^51^.

When we compared different endpoints using LASSO regression, we found a larger effect size in the reduction in IPD hospitalization than in all-cause pneumonia hospitalization. As expected, using a more specific endpoint gave estimates of larger effect size because a larger fraction of the measured outcome was caused by the pathogen of interest, which is consistent with prior studies, in which different methods were used for the PCV impact estimation^16,17,23,52^.

Our results showed that applying LASSO regression to pneumonia hospitalization data was sensitive to removing “bronchitis and bronchiolitis” from the pool of control variables subject to selection by LASSO. One possible explanation for this observation is a potential violation of our assumption that bronchitis and bronchiolitis hospitalization was not impacted by PCV. Bruhn et al.^24^ and Jimbo Sotomayor et al.^37^ highlighted that including bronchitis and bronchiolitis hospitalization can be important for the accurate prediction of pneumonia hospitalization, especially in young children, due to its association with respiratory syncytial virus (RSV) infections. Of note, the fraction of bronchitis and bronchiolitis hospitalization caused by the pneumococcus and the prevalence of RSV differ by age groups^53–56^; therefore, it is important to consider the potentially different pathogen-pathogen dynamics in different age groups when estimating vaccine impact.

There are a few major limitations in our study that should be considered. First, the simulated data were generated based on the time series of ten or fewer causes of hospitalization, and LASSO tends to perform well in situations where a few variables predict outcome because of its property to eliminate variables by shrinking their coefficients to zero. Therefore, the simulation scenarios in our study may favor LASSO regression. Nevertheless, it is possible that pneumonia hospitalization can be predicted by a few control conditions given its relatively strong seasonality and well-established etiology. Second, we extracted 95% PI from the Poisson distribution of the predicted values of the outcome during the evaluation period, and, as a result, only the uncertainty around the Poisson distribution was considered. The narrow 95% PI may therefore be over optimistic and cannot be compared to the 95% CI in SC^23,25^ or STL+PCA^24^, which consider the uncertainty of the included parameters. Third, monthly case counts lower than 10 were masked in the US due to privacy reasons, and we imputed these masked values by randomly selecting a number between 0 and 9. It is less likely to pose problems to the primary endpoint analysis because pneumonia hospitalization case counts in all age groups were high (in the scale of 100 to 10,000). In contrast, more specific endpoints such as IPD and pneumococcal pneumonia had lower hospitalization counts in younger age groups, although information from the trend was retained because the masked value had a definite range (less than 10). Lastly, using the predicted counterfactual based on pre-vaccine period data to infer vaccine impact assumes the relationship between the control conditions and the outcome remained the same before and after PCV introduction. Therefore, if the relationship between the control conditions and the outcome changed near the time point of vaccine introduction, the prediction performance of LASSO would be impacted; however, we believe it is rare for the relationship between all of the control conditions and the outcome to be altered at the same time. An exception may be the scenario of a vaccine introduced to mitigate the effects of a disease that has a very strong impact on lifestyle, mortality, and the healthcare system capacity, as was seen during the COVID-19 pandemic, for example.

Despite the aforementioned limitations, our study presents a novel approach for counterfactual prediction to serve the goal of vaccine impact inference. The validation using simulated data under a variety of scenarios and an application using epidemiological data in four countries in this study demonstrated LASSO regression’s ability to estimate PCV impact. Given its stable performance shown in this study and ease of implementation, we argue that LASSO regression can be useful to assess the impact of other vaccines and ultimately help process epidemiological data to inform health policy making.

## Data Availability

Simulation data used in this study can be obtained using authors' R code (currently available upon request, will be made publicly available). Hospitalization data used in this study have been previously published by Bruhn, Christian AW, et al. "Estimating the population-level impact of vaccines using synthetic controls." Proceedings of the National Academy of Sciences 114.7 (2017): 1524-1529, and can be obtained from the GitHub database

https://github.com/weinbergerlab/synthetic-control/

## Acknowledgements

We would like to thank Daniel M. Weinberger for making the data available and offering insightful comments, and Iris Artin for offering help with the use of the package “InterventionEvaluatR”. We also thank Annette Aigner, Madlen Schranz, Elizabeth Goult, and Laura Barrero for their helpful discussions.

