## Appendices for "Using LASSO regression to estimate the population-level impact of pneumococcal conjugate vaccines"

#### Table of Contents

Appendix 1. The input variables

Appendix 2. Outcome simulation

Appendix 3: Variable selection by LASSO methods

Appendix 4. Other results in the simulation study (10 causal control variables, sparse data)

Appendix 5. Sensitivity test after removing bronchitis and bronchiolitis

#### Appendix 1. The input variables

We simulated the outcome based on a combination of control variables, one seasonal variable, and an offset. First, we used 5 randomly selected control variables to generate the outcome, we repeated this process five times, each time randomly selecting a different set of 5 control variables. Then, we increased the number of causal control variables to 10 and again, we repeated the process five times. Table S1 shows the complete list of variables, where highlighted in grey are the control variables that were ever selected to simulate the outcome. In the non-causal framework, we simulated the outcome using 3 control variables, and then removed the 3 causal control variables alongside the control variables that belonged to the same chapter under the International Classification of Diseases 10th revision (ICD-10). In Table S1, the control variables that were ever selected to simulate the outcome in the non-causal framework are marked with "*" and the control variables that were then removed are marked with "†".

Table S1. The input variables in the simulation study

| Variable Type | Variable code | Description | Exclusion |
|---|---|---|---|
| Offset | ach_noj | All non-respiratory hospitalizations | J00—99, F and O chapters |
| Control variables | A10—B99 | Non-pneumococcal infections | A40, A49, B95 |
| | A41 | Non-pneumococcal septicemia | |
| | B20—24 | HIV | |
| | B34 | Viral infections of unspecified sites | |
| | C00—D48*† | Neoplasm | |
| | D50—D89 | Hematological conditions | |
| | E00—99 | Endocrinological and nutritional conditions, metabolic disorders | |
| | E10—14*† | Diabetes | |
| | E40—46† | Malnutrition | |
| | G00—99 | Neurological conditions | G00—04 |
| | H00—99 | Eye and ear conditions | H10, 65, 66 |
| | I00—99 | Cardiovascular conditions | |
| | I60—64 | Stroke | |
| | J20—22*† | Bronchitis and bronchiolitis | |
| | K00—99*† | Gastrointestinal conditions | |
| | K35† | Appendicitis | |
| | K80† | Cholelithiasis | |
| | L00—99 | Dermatological conditions | |
| | M00—99 | Musculoskeletal conditions | |
| | N00—99† | Gynecological conditions | |
| | N39*† | Urinary tract infection | |
| | P00—99 | Neonatal conditions | |
| | P05—07 | Premature delivery and low birth weight | |
| | Q00—99 | Congenital or developmental conditions | |
| | R00—99 | Symptoms and signs, abnormal clinical or lab findings without diagnosis | |
| | S00—T99 | Injury, poisoning, and conditions due to external causes | |
| | U00—99 | Codes for special purposes | |
| | V00—Y99 | Accidents and trauma | |
| | Z00—99*† | Health examinations and disease screening | |
| Seasonal variables | Cosine waves | $\cos(n * (2\pi/12) * t)$, $1 \leq n \leq 6$ | |
| | Sine waves | $\sin(n * (2\pi/12) * t)$, $1 \leq n \leq 5$ <br>($t$ is measured in months) | |

### Appendix 2. Outcome simulation

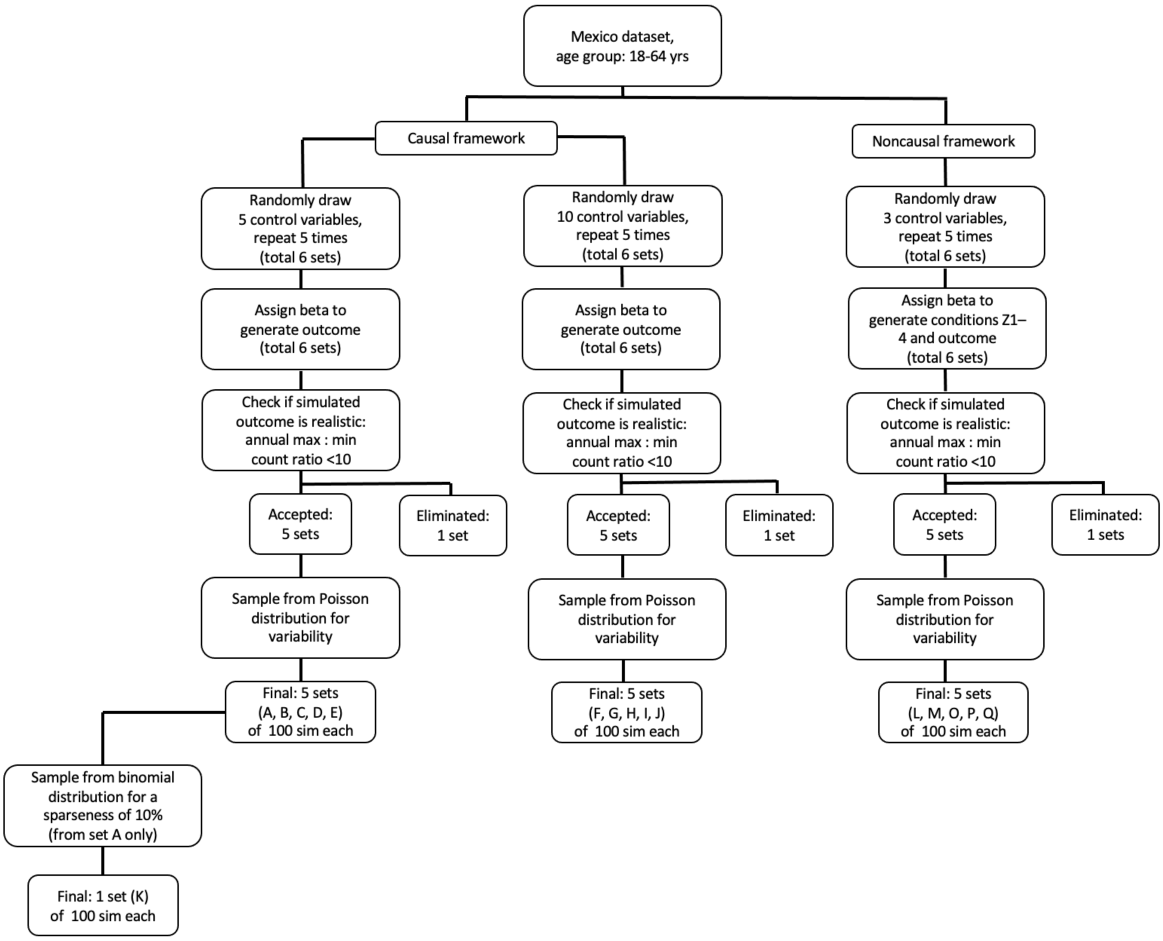


Figure S1. A flow-chart illustrating the procedure of outcome simulation.

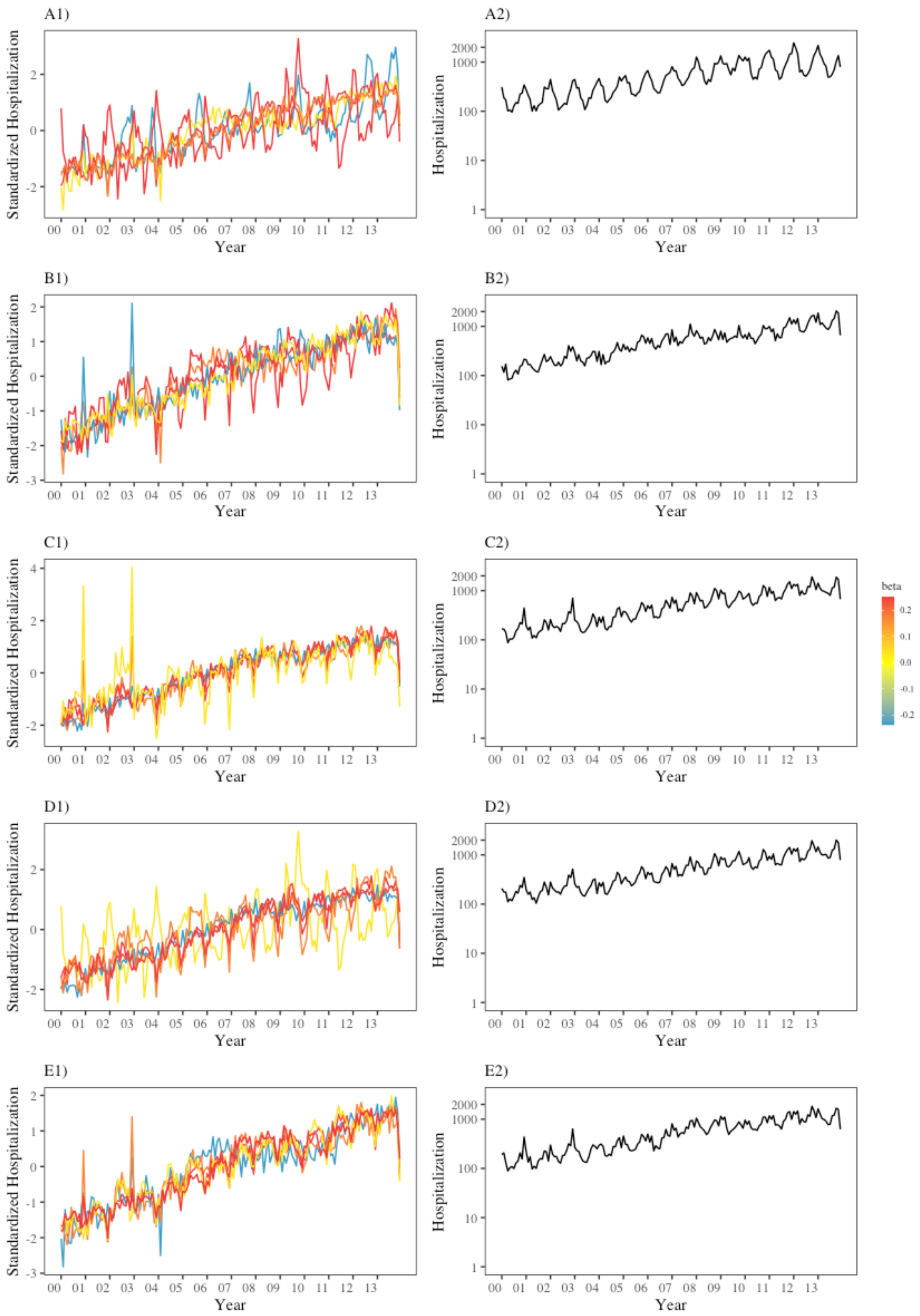


Figure S2. Time series of the five control variables (log-transformed, standardized) selected for outcome simulation and the simulated outcome in sets A to E.

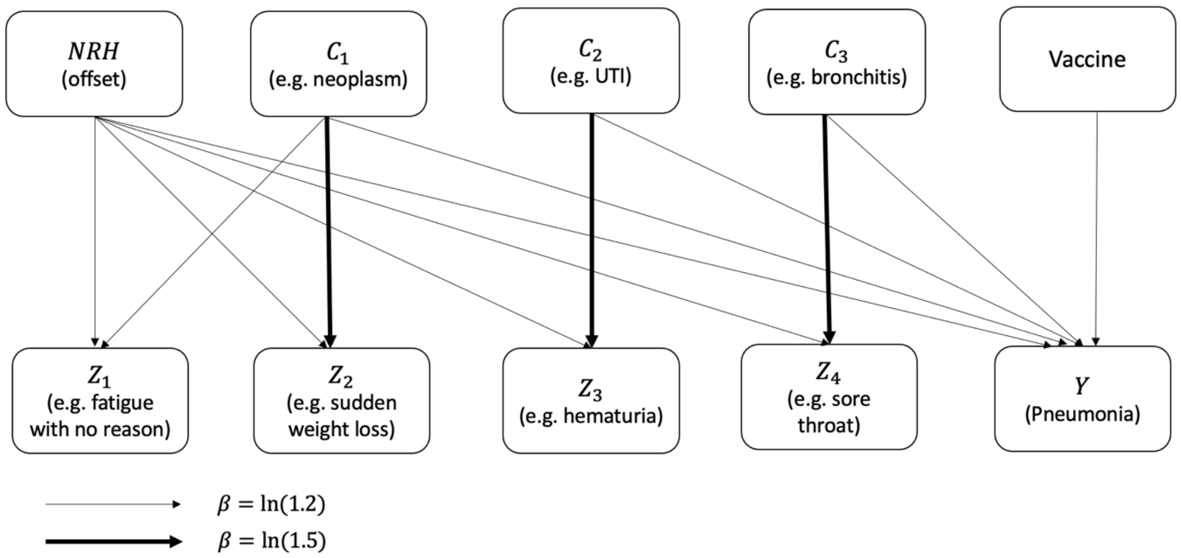


Figure S3. Directed Acyclic Graph (DAG) for outcome simulation under the non-causal framework. The causal relationships represented in this DAG depict the relationships between the control variables ($Z_1$, $Z_2$, $Z_3$, $Z_4$) and the outcome ($Y$) simulated under the non-causal framework in this study. The thickness of the arrow represented the magnitude of beta coefficient assigned to the causes ($C_1$, $C_2$, $C_3$). The causes (marked with "*" in Table S1) together with their associated control variables (marked with "†" in Table S1) were then removed from the list of control variables for model testing.

#### Appendix 3. Variable selection by LASSO methods

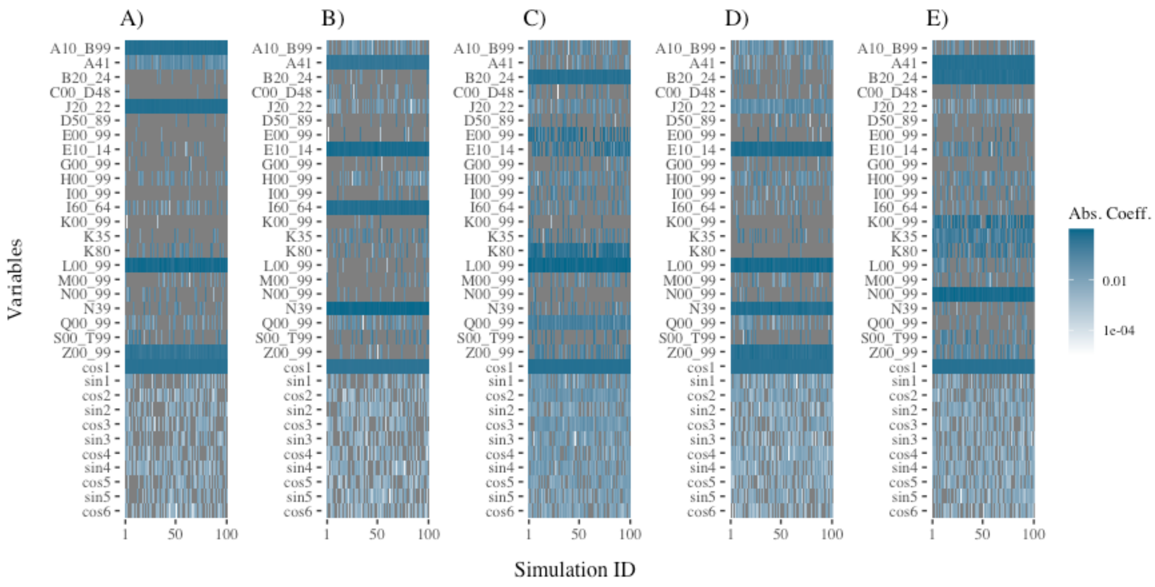


Figure S4. Seasonal and control variables selected by LASSO-SU in for simulation 1 to 100 in five scenarios, each using a different set of five causal control variables and one seasonal variable (which remained in data set) to simulate the outcome.

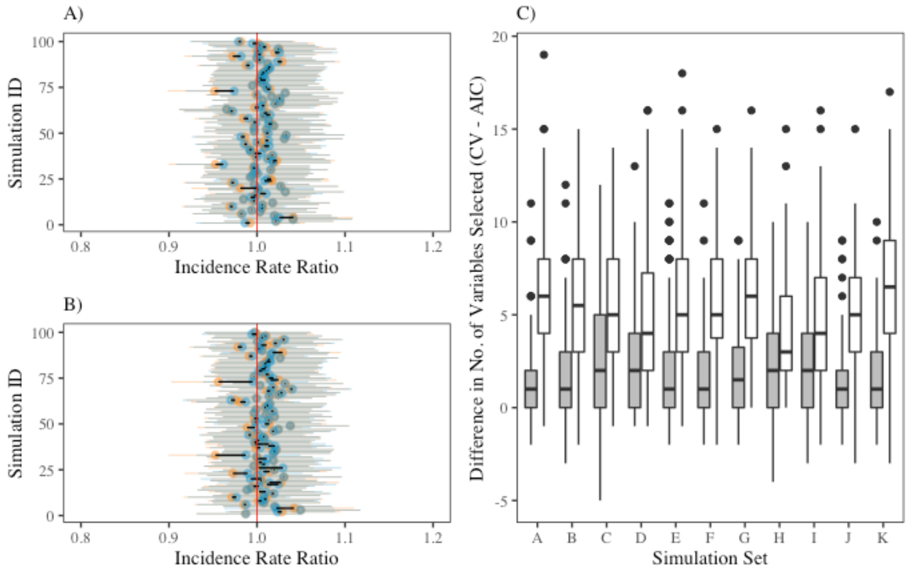


Figure S5. The difference in cross-validation (CV) vs. Akaike Information Criterion (AIC) model selection in LASSO-SF and LASSO-SU.

#### Appendix 4. Other results in the simulation study

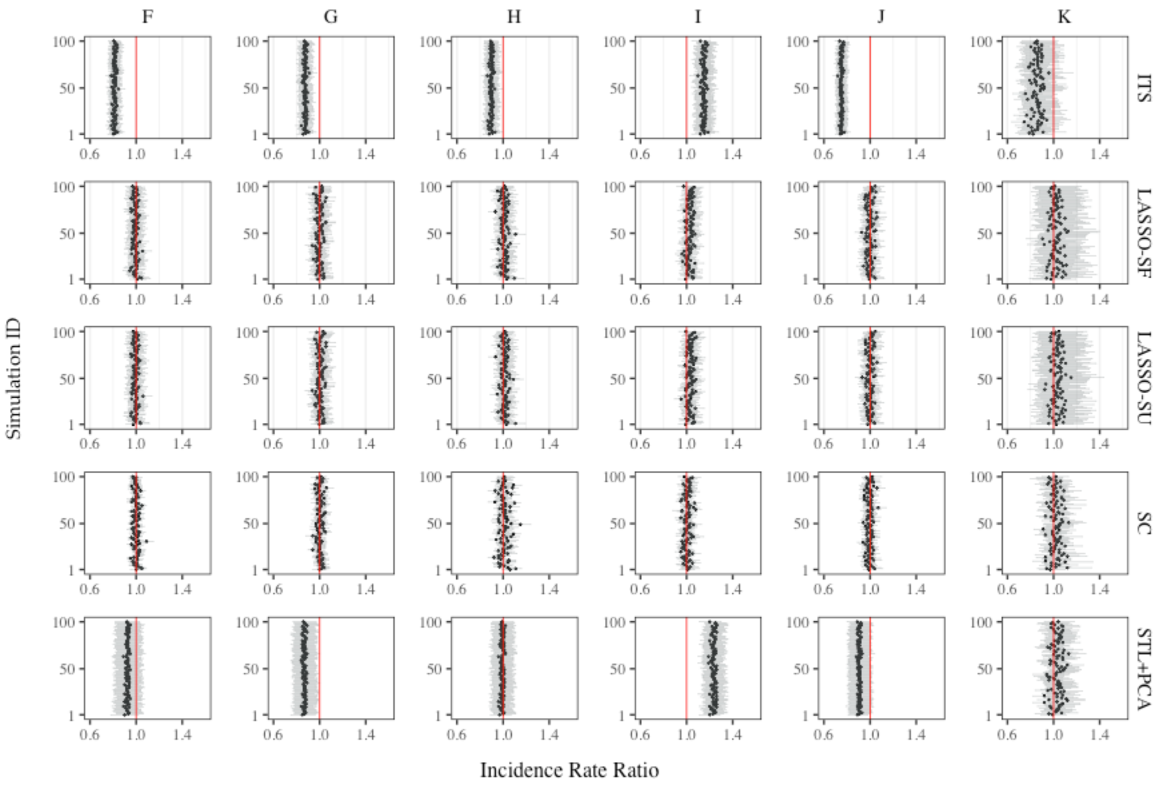


Figure S6. Results from simulation sets F to K (10 causal control var & sparse).

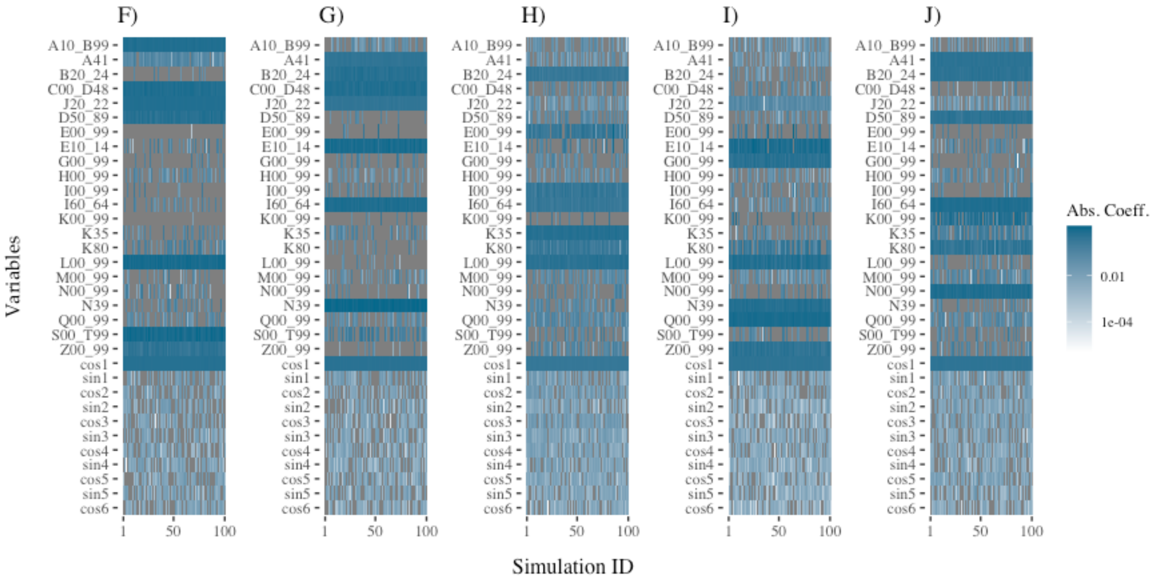


Figure S7. Variables selected by LASSO-SU in simulation sets F to J (10 causal control variables).

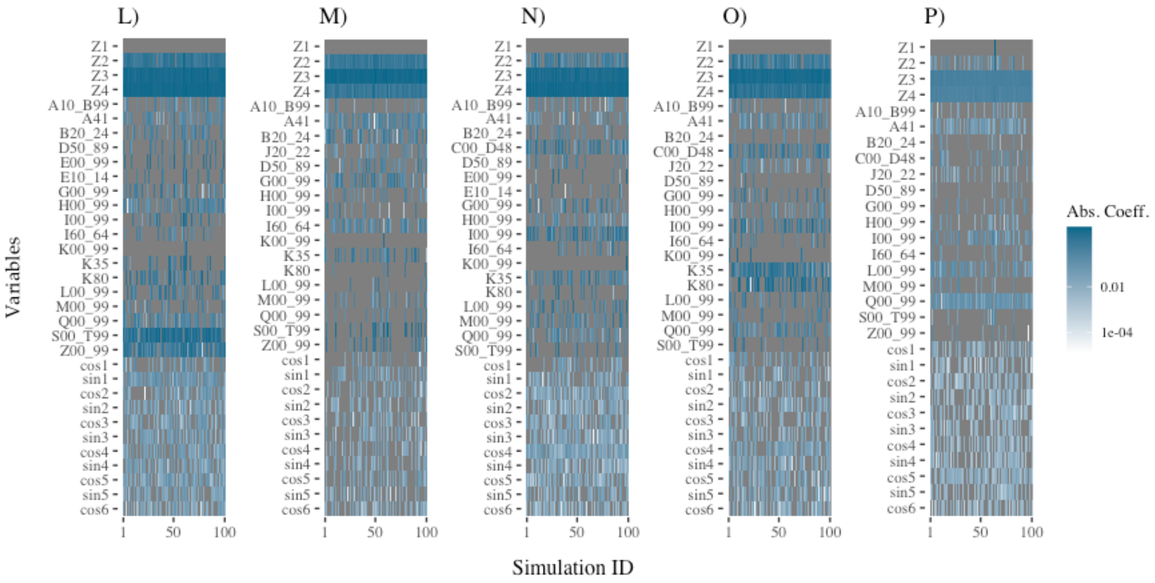


Figure S8. Seasonal and control variables selected by LASSO-SU in for simulation 1 to 100 in five scenarios, each using a different set of three causal control variables (which were then removed from data set) to simulate the outcome.

#### Appendix 5. Sensitivity test after removing bronchitis and bronchiolitis

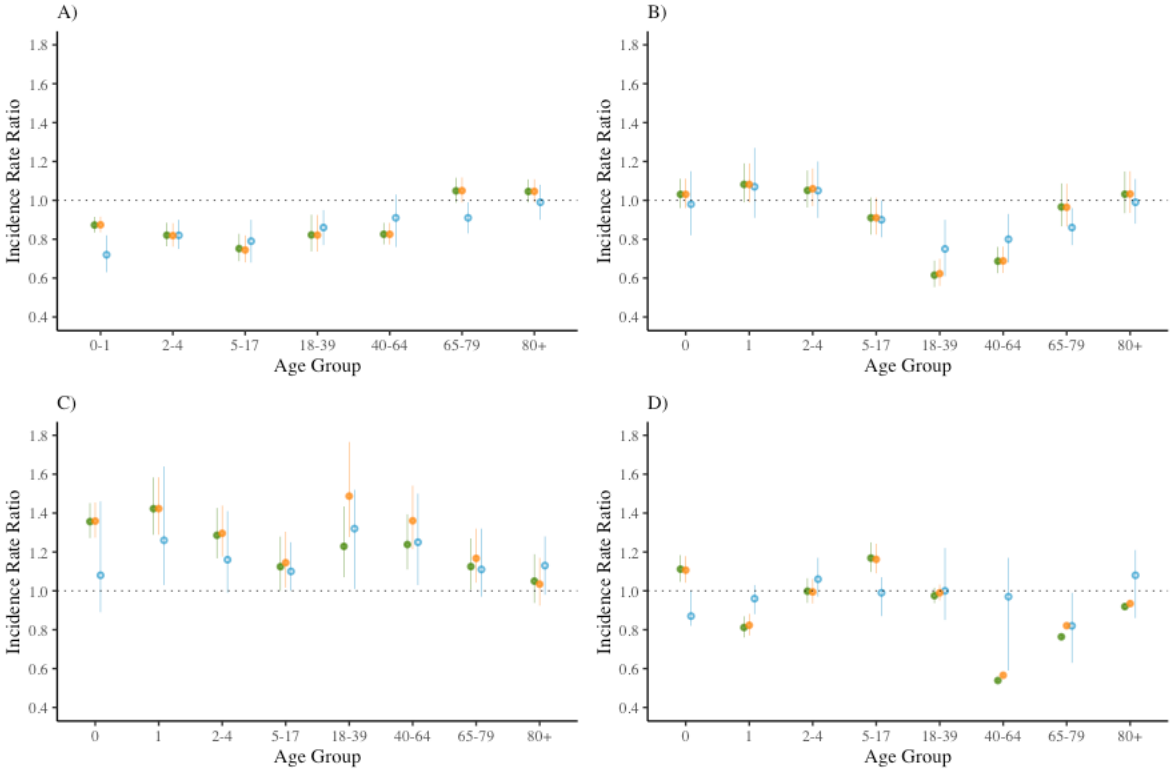


Figure S9. Age-group-specific incidence risk ratios (IRR) for all-cause pneumonia in four countries after removing “bronchitis and bronchiolitis”, estimated by two LASSO methods and SC.
